# Reporting the life tracks of confirmed cases can effective prevent and control the COVID-19 outbreak in China

**DOI:** 10.1101/2020.04.01.20050450

**Authors:** Jie Zhang, Tianjing Wang, Jiaqi Wang, Jingjing Chen, Hongwei Yan, Lin Sun

## Abstract

**Background:** Since late December 2019, a novel coronavirus (COVID-19) has emerged in Wuhan and rapidly spread throughout China. Fears were raised higher, effective policies for prevention and control were concerned.

**Methods:** Till March 3, 91273 confirmed COVID-19 cases were included in. Using Mann-Whitney U test, the provinces that reported the life tracks of confirmed cases to public had lower increased in daily new confirmed cases.

**Results:** Compared with the paired province, Tianjin, Jilin, Gansu, Shanxi, Hainan and Guizhou had significant differences in the number of new confirmed cases and have lower mean rank (P<0.05). Shanghai had a lower mean rank (P=0.175) but no significant difference. Besides, the successful prevention and control work need other effective strategies, such as real time media coverage, isolating the suspected cases or close contacts, delaying the return of work, closing schools, wearing facial masks, and disinfecting the communities, et.al.

**Conclusions:** Strategies must be adjusted in real time according to the epidemic. Reporting the life tracks of confirmed cases was an effective way to control the epidemic. They may be suggestions to other counties with an outbreak of COVID-19.

## Introduction

The current outbreak of 2019 novel coronavirus (COVID-19) was originated from Wuhan/Hubei province in central China. Just in merely a month, it fast spread across other provinces or cities(1, 2). After two months, other countries, such as Korean, Japan, Iran and Italy had severe situations. It has become a global health concern(3).

Evidence pointed out there was a strong person-to-person transmission in hospital and family settings(4-8). Despite of lockdown of the Wuhan city since 23 January 2020, the massive human movement during the period of Chinese traditional new year may have fueled the spread of the disease. As we know, general rules for the prevention and control of infectious diseases are to cut off the routes of transmission, to control the source of infection, and to protect vulnerable populations. To date, there is no effective antiviral treatment or vaccine specifically designed for this virus, which is the seventh member of enveloped RNA coronavirus(2). In this situation, some non-pharmaceutical interventions, closing the public gathering places, waring facial masks and social distancing could slow the spread of the disease. However, just one intervention, may be ineffective for this disease. So, effective public health policies, issued or executed by government, are needed for the disease preventive and control. Based on the experience of SARS (Severe Acute Respiratory Syndrome) prevention and control in 2003(9-12), the Chinese government has carried out a rapid and effective prevention and control(13).

In this study, by collecting the published data from the Health Commission of 23 provinces, 5 autonomous regions, 4 municipalities directly under the Central Government and 2 special administrative regions, we compared the effect of epidemic prevent and control for each paired provinces or cities which were based on the population density of the administrative population and the administrative area.

## Materials and Methods

### Data collection

Public data on official websites of Health Commission of 23 provinces, 5 autonomous regions, 4 municipalities directly under the Central Government and 2 special administrative regions were retrieved manually. The published date, the number of confirmed COVID-19 cases and the detailed level of data description including symptoms, transmission routes, etc. were recorded. From January 21^st^ to March 3^rd^, 80303 cases of China were included in. All information above was collected by two persons independently.

### Population density

We calculated the population density of the administrative population and the administrative area. The population of administrative regions in various provinces and cities were based on the China Statistical Yearbook 2019 (http://www.stats.gov.cn/tjsj/ndsj/2019/indexch.htm), and the area of administrative regions in various provinces and cities were based on the China’s second national land survey(14).

### Statistical analysis

Time distribution of confirmed cases with the public health policies were compared between different areas in China by line charts. For testing hypothesis that whether the report on life tracks of confirmed cases to public is useful for the epidemic, the number of confirmed cases were compared by Mann-Whitney U test. A pair of provinces or cities were matched if they had similar population densities. A two-tailed P < 0.05 was considered as statistically significant. ALL statistical analyses were done using the SPSS software, version 20.0 (IBM Corporation, Armonk, NY).

## Results

### Situation reports of COVID-19 in various provinces of China

Till March 3^rd^, there were 80303 confirmed cases, 2947 deaths and 47281 recurred cases in domestic (Table 1). The top five provinces had more than 1000 cases. Hubei province had 67214 confirmed cases. Guangdong, Henan, Zhejiang, Hunan, and Anhui had 1350, 1272, 1213 and 1018 confirmed cases respectively. The existing confirmed cases in Hubei, Guangdong, Shandong, Sichuan and Zhejiang were 28216, 259, 250, 146 and 130 respectively. Xizang and Qinghai had no existing confirmed cases.

**Table 1.** Existing cases, confirmed cases, deaths and recurred cases of COVID-19 acute respiratory disease reported by provinces, regions and cities in China, Data as 03 March 2020. **Abbreviations:** COVID-19: a novel coronavirus outbreak in Wuhan city in 2019

| Area | Existing cases | Confirmed cases | Deaths | Recurred cases |
| --- | --- | --- | --- | --- |
| Hubei | 28179 | 67217 | 2835 | 36203 |
| Wuhan | 24111 | 49426 | 2252 | 23063 |
| Other cities | 4068 | 17791 | 583 | 13140 |
| Guangdong | 242 | 1350 | 7 | 1101 |
| Henan | 19 | 1272 | 22 | 1231 |
| Zhejiang | 120 | 1213 | 1 | 1092 |
| Hunan | 113 | 1018 | 4 | 901 |
| Anhui | 56 | 990 | 6 | 928 |
| Jiangxi | 64 | 935 | 1 | 870 |
| Shandong | 241 | 758 | 6 | 511 |
| Jiangsu | 75 | 631 | 0 | 556 |
| Chongqing | 101 | 576 | 6 | 469 |
| Sichuan | 141 | 538 | 3 | 394 |
| Heilongjiang | 106 | 480 | 13 | 361 |
| Beijing | 118 | 414 | 8 | 288 |
| Shanghai | 41 | 338 | 3 | 294 |
| Hebei | 12 | 318 | 6 | 300 |
| Fujian | 36 | 296 | 1 | 259 |
| Guangxi | 48 | 252 | 2 | 202 |
| Shaanxi | 28 | 245 | 1 | 216 |
| Yunnan | 3 | 174 | 2 | 169 |
| Hainan | 12 | 168 | 5 | 151 |
| Guizhou | 30 | 146 | 2 | 114 |
| Tianjin | 15 | 136 | 3 | 118 |
| Shanxi | 13 | 133 | 0 | 120 |
| Liaoning | 20 | 125 | 1 | 104 |
| Hong Kong SAR | 62 | 100 | 2 | 36 |
| Jilin | 9 | 93 | 1 | 83 |
| Gansu | 4 | 91 | 2 | 85 |
| Xinjiang | 5 | 76 | 3 | 68 |
| Inner Mongolia | 15 | 75 | 1 | 59 |
| Ningxia | 5 | 74 | 0 | 69 |
| Taipei and environs | 29 | 42 | 1 | 12 |
| Qinghai | 0 | 18 | 0 | 18 |
| Macao SAR | 2 | 10 | 0 | 8 |
| Xizang | 0 | 1 | 0 | 1 |
| **Total** | **29964** | **80303** | **2948** | **47391** |

### Comparison of COVID-19 in Wuhan city, other cities in Hubei province and other provinces

Figure 1 described the trend of COVID-19 in Wuhan city, other cities in Hubei province and other provinces, and the main strategies in China. On December 1, first case was confirmed in Wuhan city. Since Jan 25^th^, the confirmed cases were increased in Wuhan city, other cities in Hubei province and other provinces, municipalities and special administrative regions. Wuhan city had the largest confirmed cases every day, then other cities in Hubei province and then other provinces and cities. On Feb. 12^th^, 15162 cases were confirmed nationwide. 15152 cases were diagnosed by clinical diagnosis criteria, including 1332 cases in Wuhan city. After Feb.20^th^, there were almost 400 confirmed cases in Wuhan city, no more than 100 cases in Hubei province (except Wuhan city), no more than 10 cases in other provinces.

**Figure 1.**
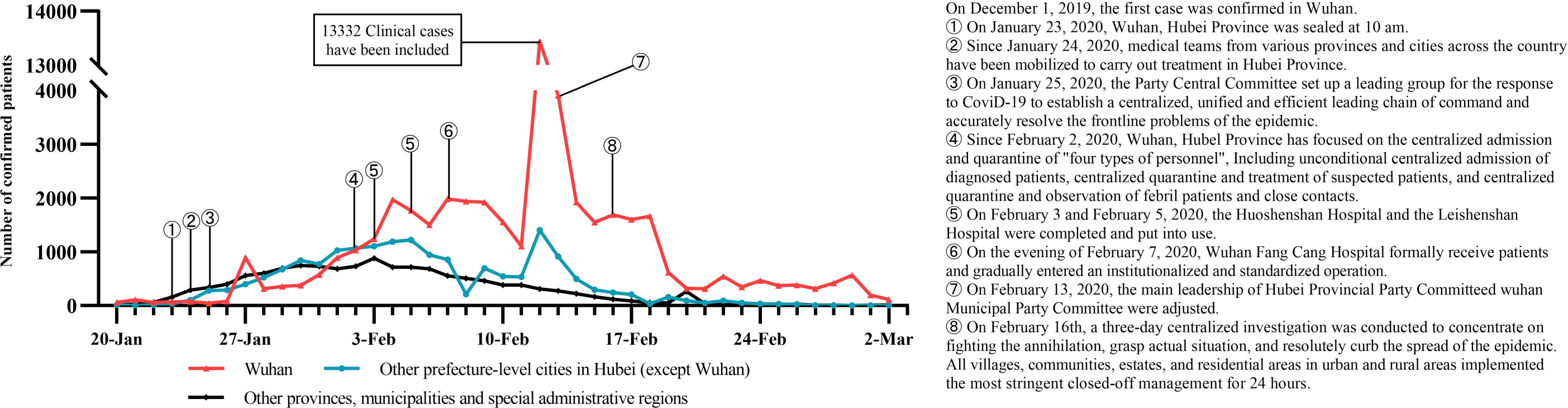
The comparison of COVID-19 confirmed cases in Wuhan city, other cities in Hubei province and other provinces. **Abbreviations:** COVID-19: a novel coronavirus outbreak in Wuhan city in 2019

### Impacts of whether reporting the life tracks of confirmed cases in different provinces

There were different levels of situation reports in various provinces in domestic. In 34 units, seven provinces or municipalities directly under the central government had published the life tracks of confirmed cases on their official websites. Tianjin, Jilin, Gansu, Shanxi and Hainan reported the detail life tracks of confirmed since Jan.25^th^, Jan.22^nd^, Jan.23^rd^, Jan.22^nd^, and Jan 22^nd^, respectively. These five units had a milder outbreak, and up to March 3^rd^, the existing cases in the five units were no more than 15. Table 1 showed they ranked behind other provinces. Shanghai and Guizhou reported the detail life tracks of confirmed cases in the middle of the epidemic since Feb.9^th^ and Feb.5^th^, respectively. Among the provinces that have never published their life tracks, other seven other provinces were matched to the seven provinces with the similar population density respectively. Then we got seven pairs of units, each pair had one unit that published the life tracks of confirmed cases and one unit did not publish such information. The comparison of daily confirmed cases and cumulative confirmed cases shown in Figure 2 and Figure 3, respectively. The epidemic was worse in provinces which had no reporting the life tracks.

**Figure 2.**
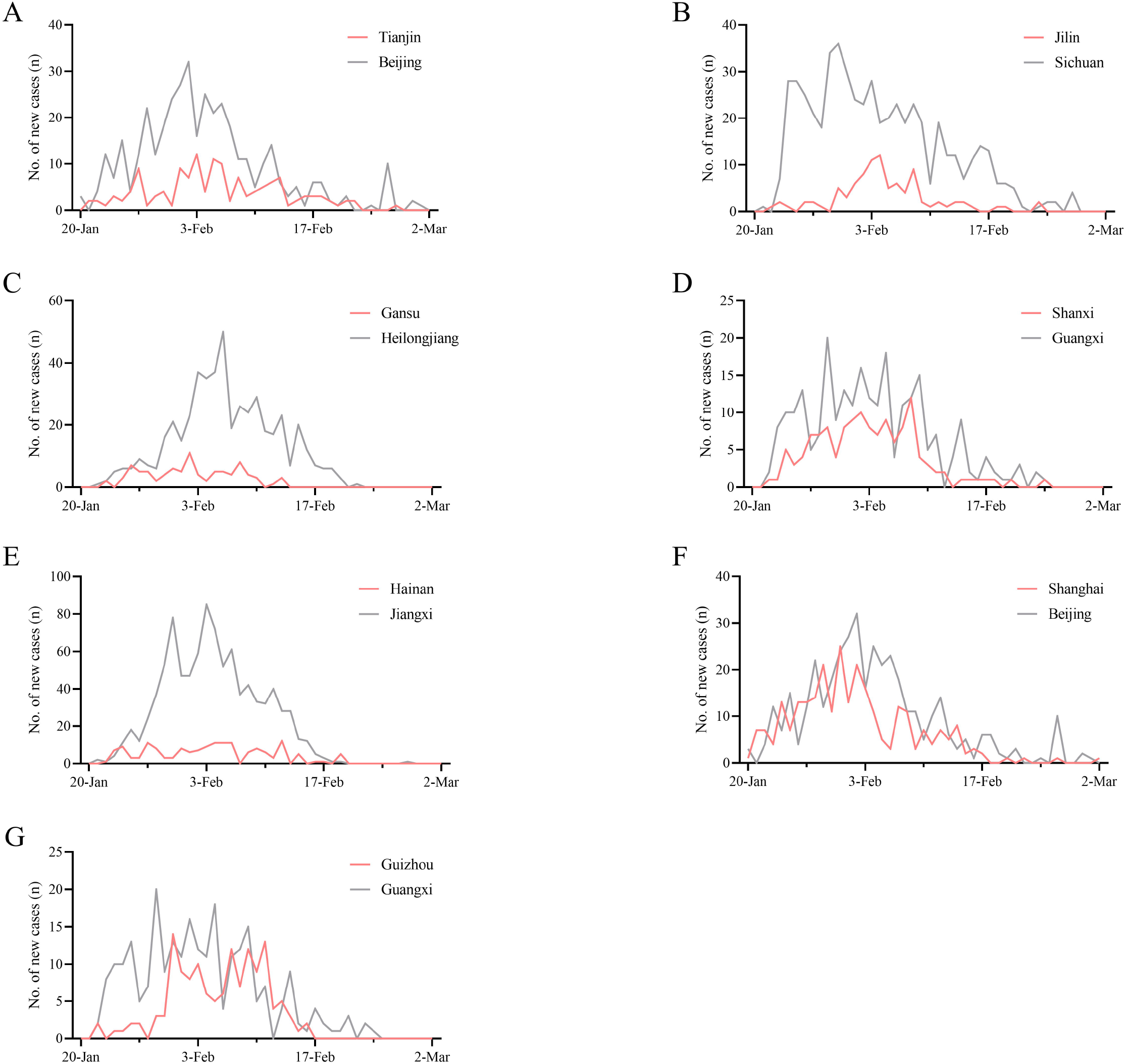
The comparison of daily confirmed cases in the provinces where the life track is published or not. (A) The comparison of Tianjin and Beijing. (B) The comparison of Jilin and Sichuan. (C) The comparison of Gansu and Heilongjiang. (D) The comparison of Shanxi and Guangxi. (E) The comparison of Hainan and Jiangxi. (F) The comparison of Shanghai and Beijing. (G) The comparison of Guizhou and Guangxi.

**Figure 3.**
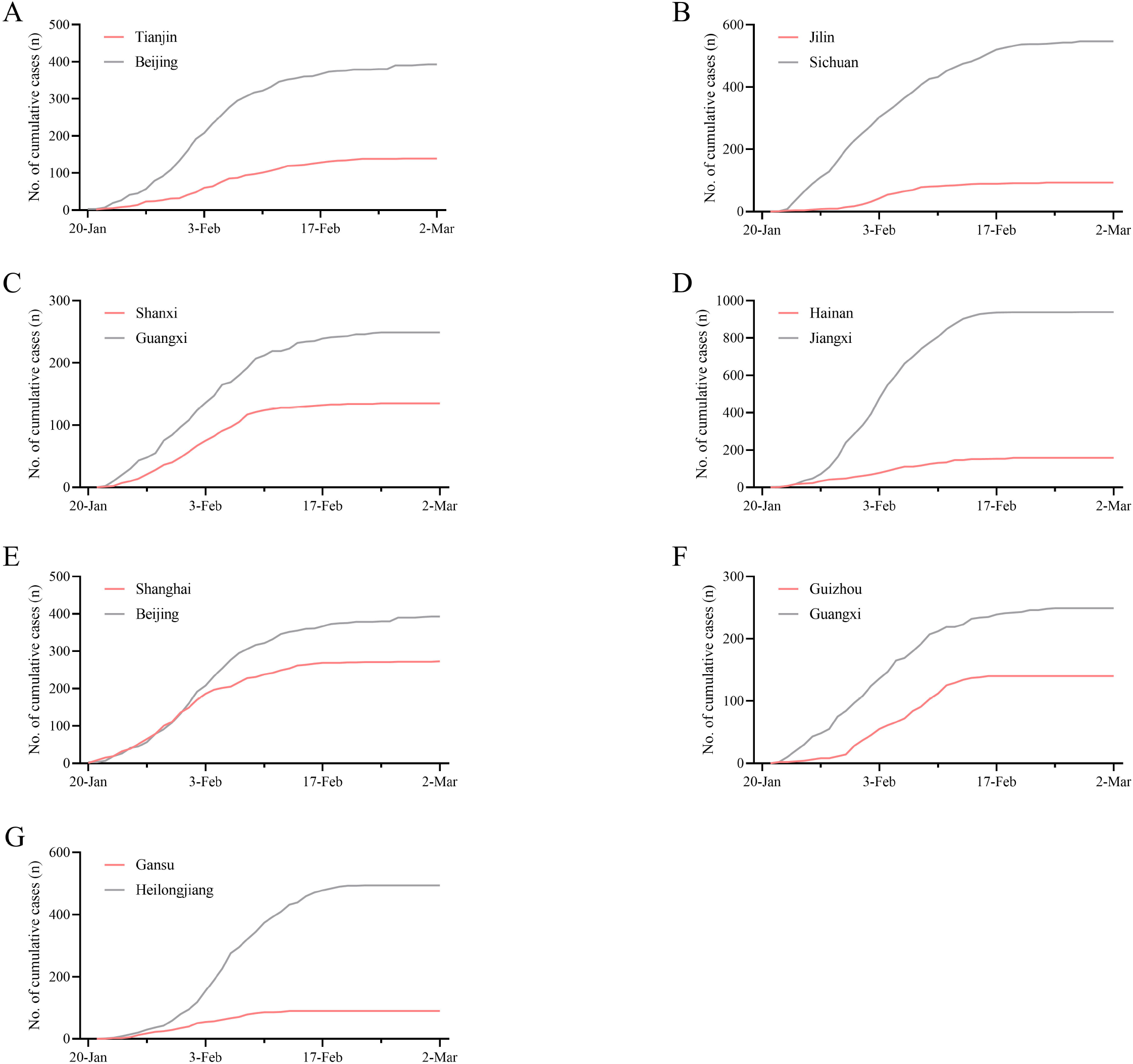
The comparison of cumulative confirmed cases in the provinces where the life track is published or not. (A) The comparison of Tianjin and Beijing. (B) The comparison of Jilin and Sichuan. (C) The comparison of Gansu and Heilongjiang. (D) The comparison of Shanxi and Guangxi. (E) The comparison of Hainan and Jiangxi. (F) The comparison of Shanghai and Beijing. (G) The comparison of Guizhou and Guangxi.

The comparison of daily confirmed cases in the provinces where the life track is published or not Through Mann-Whitney U test, it was found that the difference between the two units of six pairs was statistically significant (Table 2). Compared with the paired province, Tianjin, Jilin, Gansu, Shanxi, Hainan and Guizhou have significant differences in the number of new confirmed cases and have lower mean rank (P<0.05). Compared with the matched municipalities, Shanghai has not significant difference but has a lower mean rank (P=0.175).

**Table 2.** Comparison between pair of regions that published the life tracks of confirmed cases or not based on Mann-Whitney U Test

| Regions |  | Matched regions |  | *P* | Mean Rank |  |
| --- | --- | --- | --- | --- | --- | --- |
| Name | Population density<br>( person/square<br>kilometer ) | Name | Population density<br>( person/square<br>kilometer ) |  | Reported<br>regions | Unreported<br>regions |
| Tianjin | 1309.05 | Beijing | 1312.93 | 0.002 | 33.23 | 49.77 |
| Jilin | 141.42 | Sichuan | 171.58 | <0.001 | 29.20 | 53.80 |
| Gansu | 61.92 | Heilongjiang | 83.37 | <0.001 | 30.96 | 52.04 |
| Shanxi | 237.27 | Guangxi | 207.32 | 0.032 | 35.94 | 47.06 |
| Hainan | 265.51 | Jiangxi | 278.43 | 0.001 | 33.26 | 49.74 |
| Shanghai | 2899.87 | Beijing | 1312.93 | 0.175 | 37.95 | 45.05 |
| Guizhou | 204.43 | Guangxi | 207.32 | 0.026 | 35.76 | 47.24 |

## Discussion

In this highly mobile world, the outbreak of COVID-19 is a challenging task to China. In particular, there is a strong person to person transmission(5) and the treatment and prevention options are limited. To control the novel pathogen could be a challenging task for China. Based on the prevention and control for MERS (Middle East Respiratory Syndrome)(15, 16) or SARS (Severe Acute Respiratory Syndrome)(9-12, 17-20), Chinese government had issued some effective policies or proposed some precautions in case detection and management, vulnerable population protection, and transmission routes cutting off. To date, these measures are very effective to prevent and control the spread of COVID-19.

An outbreak requires attention in advance to clarity, collaboration, communication, coordination and capacity(21). During the outbreak of SARS, policies were issued by Chinese government. Such as media reporting in real time, built Xiaotangshan hospital for confirmed disease, and published the life tracks of confirmed cases, which may make people were more alert. The spread of SARS in 2003, also brought to light substantial weaknesses in the country’s public-health system. After the SARS epidemic was brought under control, the government increased its commitment and leadership to tackle public-health problems and, among other efforts, increased public-health funding, revised laws that concerned the control of infectious diseases, implemented the world’s largest internet-based disease reporting system, and started a program to rebuild local public-health facilities(22).

For the outbreak of epidemic in the whole country, the prevention and control system should be changed timely. Different levels of cases should be treated differently. Case detection and management, vulnerable population protection, and transmission routes cutting off are needed to decrease the rate of infection. The monitoring of the epidemic by Health Commission is necessary. Real time media are needed to attract the people’s attention. In this study, we found the provinces that reported the life tracks for confirmed cases had mild epidemic and less confirmed cases (Figure 2 and Figure 3). Compared with the paired province, Tianjin, Jilin, Gansu, Shanxi, Hainan and Guizhou have significant differences in the number of new confirmed cases and have lower mean rank (P<0.05, table 2). Compared with the matched municipalities, Shanghai has not significant difference but has a lower mean rank (P=0.175). There may be two reasons for this result. Firstly, the population densities were the best matched, but there was a big gap between Beijing and Shanghai. Secondly, the reports on the life tracks for confirmed cases in Shanghai began later not at the beginning.

From our opinion, there were two advantages for the epidemic prevention and control for COVID-19. Firstly, in this period, most people stay at home. They always to read the daily media report, and talk about the number of confirmed cases in Wechat or other media social platforms. They can be alert if they find the confirmed case living near their house. Or they may avoid to go the places where the confirmed cases stayed by. So, for the public, reporting the life tracks of confirmed cases is an effective way to help them keep away from the ‘infectious’ things or the places. This may good alert for protect the public. Secondly, for tracking the detail information of life tracks would need more time and work for some of government administration departments. Whether the local governments are willing to put in the effort and time in it, to some extent, may showed how meticulous and important the local governments were in their work.

Although the report for the life tracks of confirmed cases had some controversial, it was proved that this kind of information was effective and feasible in our country. To date, except Wuhan city and other cities in Hubei province, we found the number of new COVID-19 cases increased slowly, even in some provinces, there were no new cases increased these days. Nine provinces had no more than 10 cases, and Xizang and Qinghai provinces had no confirmed case now. Even Wuhan city had a decreased number of confirmed cases every day. The information showed a controlled trend for this epidemic outbreak.

The epidemic was under control in China. Still, there were some limitations in our study. Firstly, the outbreak in China is not over yet. There may be another peak of infection rate with the resumption of work. Secondly. Wuhan city still had a severe situation now. Anyway, the number of daily confirmed cases were dropped sharply which showed the effective of epidemic prevention and control.

## Conclusion

COVID-19, with a strong person-to-person transmission made a pandemic in China and outbreak in Korean, Iran, Italy and Japan. Along with the decreased number of new confirmed cases and the increased number of recovered cases, there might be a turning corner of COVID-19 in China. This can be attributed to the success of a serial of emergence public measures and the confidence of the whole nation, which may be useful for the epidemic prevention and control in other counties. However, it remains a challenging task to improve the mechanism for major epidemic prevention and control and the national public health emergency management system.

## Data Availability

Public data on official websites of Health Commission of 23 provinces, 5 autonomous regions, 4 municipalities directly under the Central Government and 2 special administrative regions were retrieved manually. The published date, the number of confirmed COVID-19 cases and the detailed level of data description including symptoms, transmission routes, etc. were recorded. From January 21st to March 3rd, 80303 cases of China were included in. All information above was collected by two persons independently.

## Author contributions

JW and TW analyzed the data, JZ wrote the manuscript. JW, TW, JZ, LS, HY and JC collected the data, LS and JZ designed the research and monitored the quality of the analysis.

## Conflict of interest

The authors declare that they have no conflict of interest.

## Notes

### Competing Interest Statement

The authors have declared no competing interest.

### Funding Statement

none

